# Leukocyte Count Is Better than LDL-C as Predictor of Novel Carotid Atherosclerosis

**DOI:** 10.1101/2025.01.14.25320572

**Authors:** Y Li, H Cao, L Ding, T G Naren, Q Q Zhang, Z Wang

## Abstract

**Background:** This continuous retrospective cohort study aims to(1)screen the risk factors and cut-off values of initial occurrence of carotid atherosclerosis(CAS) and(2)identify whether the pathological procession of CAS is from carotid intima-media thickening(C-IMT) to carotid plaques CAP).

**Methods:** Between 2015 and 2024, the characteristics were recorded at three time points, which were the meaningful time point for the first new appearance of CAS or not, the baseline time point for the previous closest normal carotid status, and the validated time point for the first confirming the meaningful results. Statistics analyses, including student’s t test, Mann-Whitney U test, and Chi-square test, assessed the different results between observation group and healthy controls. Logistic regression, Cox regression, ROC curves and Kaplan-Meier analysis were used for screening the risk factors and cut-off points. Repeated-measures ANOVA was used for comparison between the groups and within each group.

**Results:** Of 3583 recruited participants, the final study analyses included 1141 individuals, there was no significant change in the proportion of C-IMT and CAP during continuous observation of a 1.04 years (P=0.561). After performed Propensity score matching for age and gender, leukocyte count 5.00*10^9^/L and low-density lipoprotein cholesterol (LDL-C) 125.1mg/dl were significantly associated with the new appearance of CAS over a 1.09 years follow-up period compared to the reference group. Leukocyte count high level group was associated with CAS (log-rank P=0.01), nevertheless LDL-C was no significant difference (log-rank P=0.055).

**Conclusions:** Middle aged adults(aged 49.6± 8.0)with leukocyte count above 5.00*10^9^/L were more likely progress CAS after an average of 1.09 years. CAS new lesions had no obvious specificity and no significant changes were found after an average of 1.04 years. This study identified early specific markers that predict the appearance of CAS in order to guide the timing of early lifestyle interventions.

## Introduction

Continuous damage to the carotid artery in the apparently healthy population has resulted in CAS. Subclinical atherosclerosis in asymptomatic individuals was independently associated with all-cause mortality^1-3^. The results of carotid ultrasound could reflect not only carotid statis, but also predict systemic arteriosclerosis^4,5^, and the ultrasound assessment of CAP, compared with that of C-IMT, had a higher diagnostic accuracy for the prediction of future CAS events^6^. However, there are no evidence to suggest whether C-IMT is the initial manifestation of carotid artery disease, whether CAP represent the progression of C-IMT.

The appearance of atherosclerosis linked to elevated levels of clinical biomarkers associated with cardiovascular injury such as markers for sympathetic activation, oxidative stress, Inflammation, thrombosis, and vascular dysfunction^7^.Lifestyle intervention and medical treatment for obese and high LDL-C levels populations could reverse CAS^8,9^. Chronic inflammation and dyslipidemia were key risk factors for acute coronary syndrome^10^. Nevertheless, what are the risk factors for CAS in the population with normal average body mass index and LDL-C levels. Thus, there is a need to identify early specific markers that predict the appearance of plaques in order to guide the timing of early lifestyle interventions.

## Materials and methods

### Study design and participants

This clinical study was a retrospective analysis that, between March 2015 and July 2024, enrolled 3583 participants (who had normal carotid artery and liver ultrasound at baseline) of the Health Management Center, Tsinghua Changgung Hospital, Beijing, China. Exclusion criteria included previous history of cardiovascular events (coronary disease, stroke), pregnancy, rheumatoid immune diseases (included rheumatoid arthritis, autoimmune hepatitis, ulcerative colitis). Of these participants, 2761 had underwent a minimum of three carotid artery and liver ultrasound examination. Excluded fatty liver disease, a total of 1141 participants followed at least once examination to validate the meaningful results(CAS or not),up to nine follow-up visits to confirm the meaningful examination. The ethics panel of Tsinghua Changgung Hospital granted permission for the research (ethics number:24634-6-01). Since the analysis only utilized unidentified data, individual consent was exempted.

### Clinical and biological evaluation

All participants were continuous recorded age, gender, smoking and drinking status, physical measurements (BMI, systolic blood pressure [SBP], diastolic blood pressure [DBP]),while past and family medical history were self-reported. After fasting for a minimum of 8 h overnight, blood samples were collected in the order of complete blood count first and then biochemical indicators. Missing values were filled in with adjacent values, the filling ratio of missing data did not exceed 1%, if there was no data from multiple follow-up visits, they were directly deleted.

### Assessment of CAS

Carotid artery measurements were done by five trained physicians who had more than 10 years of experience, utilizing Doppler ultrasound systems (Siemens Acuson X700 or Mindray DC-90Pro) with scanning in longitudinal and cross-sectional orientations from the proximal common carotid artery to the distal internal carotid artery on each side. CAS included carotid C-IMT and CAP.C-IMT was defined as carotid artery intima≥1.0mm or carotid artery bifurcation intima≥1.2mm. CAP was defined as intima-media thickness (IMT) ≥ 1.5 mm, protruded into the vascular lumen, or exhibited localized thickening greater than 50 % of the surrounding IMT^11-12^.

### Statistical analysis

Based on the meaningful carotid ultrasound results, the participants were divided into healthy controls and CAS group. Baseline characteristics were expressed as mean±SD for continuous variables or median (Q1-Q3) if marked skewness existed, and as number and percentage for categorical variable. Statistical analysis was performed using SPSS version 26.0 (SPSS Inc. Chicago, IL). These data were analyzed by student’s t test, Mann-Whitney U test, and Chi-square test. Propensity score matching was performed for age and gender were unmatched. P value less than 0.05 was considered statistically significant. Based on the results, ROC curves were drawn and calculated AUC, the cutoff points. Next, survival curves were plotted using the Kaplan-Meier analysis basing on the classification of cutoff values, and survival rates were no significant difference using the Log-Rank test. Cox regression was used for univariate and multivariate analysis. P-value less than 0.05 was used for multivariate analysis, with hazard ratio (HR) and 95% confidence interval (95% CI) displayed.

## Results

### Characteristics of subjects

According to the study flow Figure S1, a total of 1141 individuals were included in the study. In the CAS group, there were 117 individuals, aged 32-85 years at baseline. The average age of the group was 52.6±10.0 years. The prevalent comorbidities was metabolic syndrome (81.2%). The clinical characteristics and outcomes were no significant difference between CAS and healthy controls. In CAS first appearance, the CAS group included 49(41.9%) individuals with C-IMT, 46(39.3%) individuals with CAP, and 22(18.8%) individuals with both C-IMT and CAP. There was no significant difference in the allocation ratio of C-IMT and CAP between the meaningful time points and the validated time points(P=0.561). The average time interval between the baseline time points and the meaningful time points was 1.09 years (range:0.52-3.40 years). While the time between the meaningful time points and the validated time points was 1.04 years (range:0.58-4.23 years). Other statuses were shown in Table 1-2.

**Table 1.**
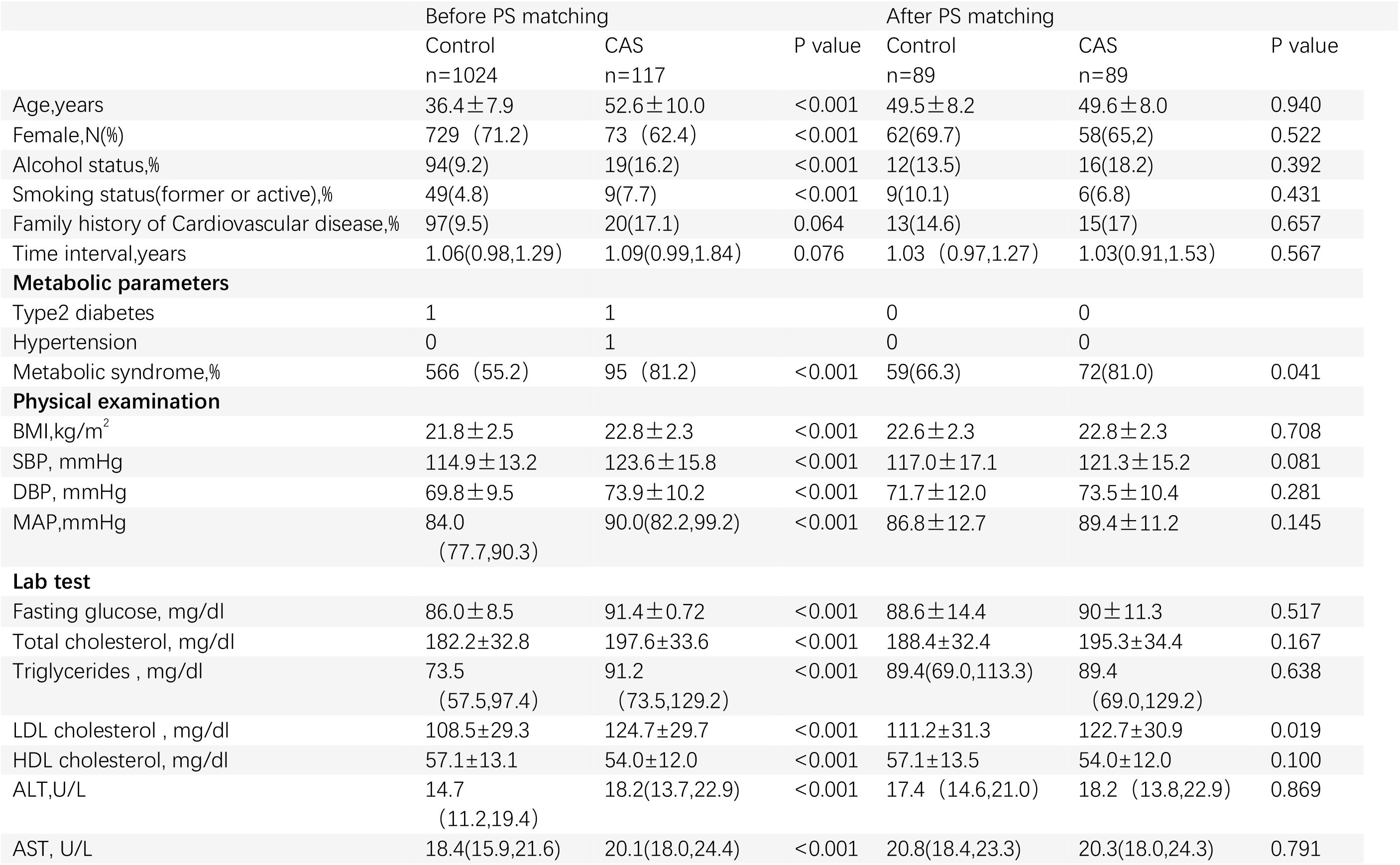
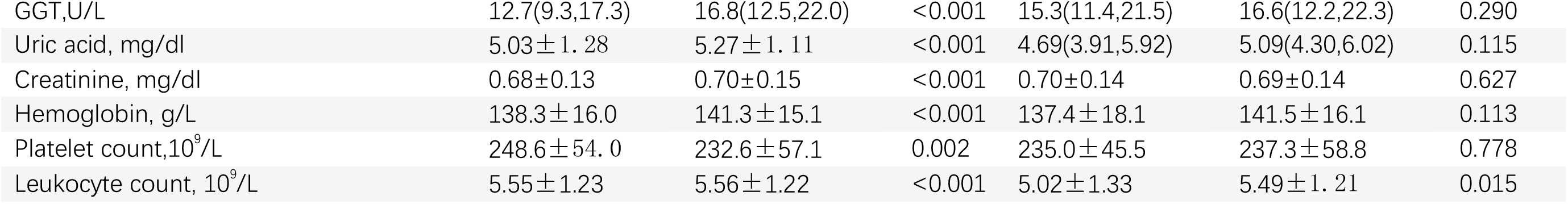
Baseline characteristics between healthy controls and CAS group before and after propensity score matching for age and gender.

**Table 2.** Characteristics of CAS participants ultrasound results before and after propensity score matching for age and sex.

|  | Before PS matching |  |  |  | After PS matching |  |  |  |
| --- | --- | --- | --- | --- | --- | --- | --- | --- |
|  | C-IMT | CAP | C-IMT+CAP | Time intervals,y | C-IMT | CAP | C-IMT+CAP | Time intervals,y |
| **Baseline time point** | 0 | 0 | 0 |  | 0 | 0 | 0 |  |
| **Meaningful time point** | 49(41.9%) | 46(39.3%) | 22 (18.8%) | 1.09(0.99,1.84) | 38 (42.7%) | 44(49.4%) | 7 (7.9%) | 1.03(0.91,1.53) |
| **Validated time point** | 44(37.6%) | 49(41.9%) | 24 (20.5%) | 1.04(0.92,1.53) | 36 (40.4%) | 39(43.8%) | 14 (15.7%) | 1.04(0.93,1.53) |

### CAS and healthy controls comparison before and after propensity score matching (PSM) for age and gender

Compared to healthy controls, CAS participants were older and had a higher percentage of males, metabolic syndrome, a higher proportion of smoking and drinking alcohol; higher levels of BMI, SBP, DBP, fasting glucose, total cholesterol, triglycerides, LDL-C, ALT, AST, GGT, uric acid level, creatinine, leukocyte count, hemoglobin, platelet count; but a lower level of HDL-C. The rates of cardiovascular family history were no significant difference between two groups. Using PSM for age and gender, CAS patients had a higher percentage of metabolic syndrome, higher levels of LDL-C and leukocyte count(Table 1).

### Factors associated with CAS using binary logistic regression and Cox proportional hazards regression analyses

For CAS patients, univariate analysis found SBP, LDL-C, leukocyte count was associated with CAS. Using binary logistic regression and Cox regression, only LDL-C, leukocyte count were associated with CAS(Table 3). Furthermore, it was found that there were significant differences in neutrophils between the control group and the CAS group, but no significant differences were observed between other types of leukocyte (Table S1).

**Table 3.** Factors associated with CAS using binary logistic regression.

|  | Univariate |  |  | Multivariate |  |  |
| --- | --- | --- | --- | --- | --- | --- |
|  | OR | 95% CI | P value | AOR | 95% CI | P value |
| **SBP** | 1.014 | 1.002-1.026 | 0.022 | 1.011 | 0.999-1.024 | 0.084 |
| **LDL-c** | 1.327 | 1.032-1.706 | 0.028 | 1.368 | 1.062-1.763 | 0.015 |
| **Leukocyte count** | 1.186 | 1.029-1.367 | 0.019 | 1.172 | 1.006-1.366 | 0.042 |

### Cutoff points of meaningful factors with CAS using ROC curves and Kaplan-Meier analysis

The Roc curves were used to analyze the predictive effect of meaningful factors on CAS patients. The areas under the curves (AUCs) for predicting CAS patients were0.622(95% CI:0.540-0.704) (P=0.005) for leukocyte count, and 0.600(95% CI:0.516-0.683)(P=0.022) for LDL-C. The AUCs were no significant difference between leukocyte count and predicted values(P=0.285), LDL-C and predicted values(P=0.285), LDL-C and leukocyte count(P=0.291).(Figure 1)

**Figure 1.**
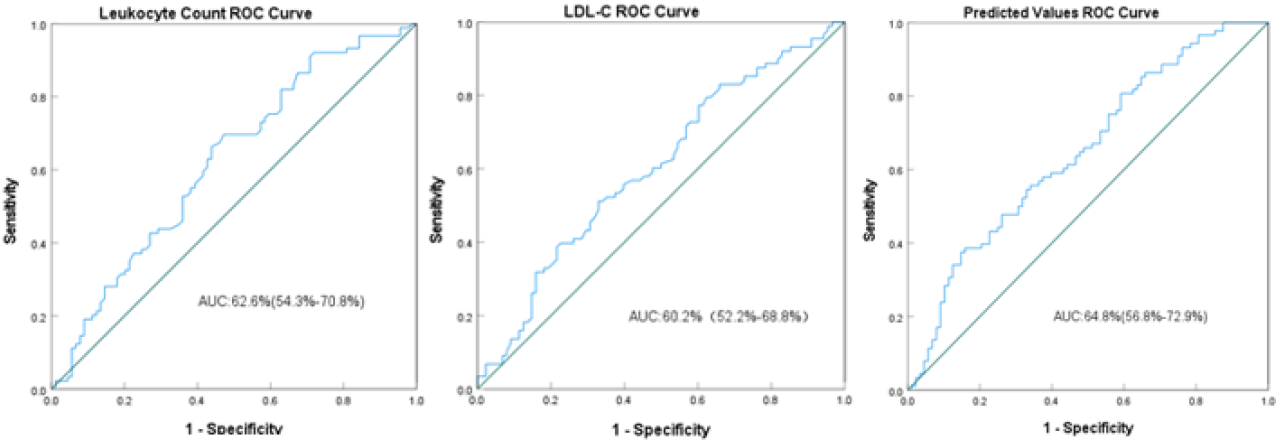
ROC curves analysis of the predictive effect of CAS progression predictors, based on the cutoff values of leukocyte count >5.00*10^9^/L and LDL-C> 125.1mg/dl respectively.↵

The cutoff points for LDL-C and leukocyte count were 125.1mg/dl and 5.00*10^9^/L respectively. Those higher than the above values were defined as high-level groups, while those below were defined as low-level groups. Kaplan Meier analysis was performed to evaluate leukocyte count and LDL-C results (Figure 2).

**Figure 2.**
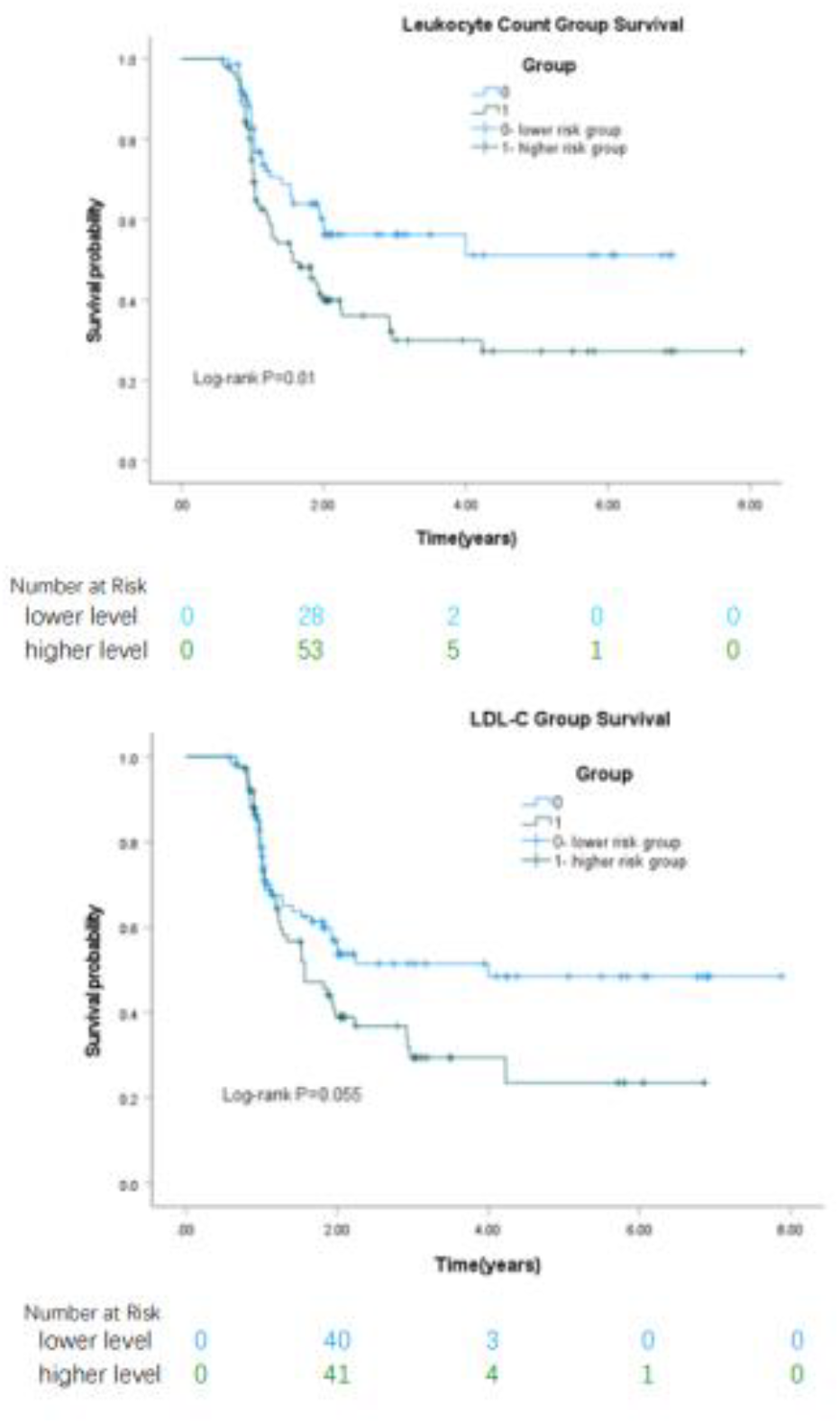
Kaplan-Meier analysis the survival rates, based on the classification of cutoff values of leukocyte count and LDL-C different levels respectively

Patients were stratified into two level groups, based on the cutoff points. Leukocyte count higher level group was associated with CAS (log-rank P=0.01), and LDL-C was no significant difference between higher risk level group and lower risk level group (log-rank p=0.055). When stratified by Leukocyte count, CAS patients and healthy participants in the low-risk were 37.5% and 62.5%, while in high-risk were 60.2% and 39.8%. Compared with the lower risk level group, the leukocyte count higher level group had significantly higher risks of CAS progression (hazard ratio(HR),1.766 :95%CI,1.138-2.743).

Increasingly, besides neutrophils, the other leukocyte subtypes were no significant differences(P>0.05). As the classification of neutrophils cutoff point(2.63*10^9^/L), there was no significant between neutrophils higher risk level group and lower risk level group (log-rank p=0.060). Therefore, the leukocyte subtypes were no significant with the first appearance of CAS.

### Examining difference within CAS group and between groups using Repeated-measures ANOVA

Elevated leukocyte count only occurred before the onset of CAS, did not increase after the appearance of CAS. In CAS group, the leukocyte counts at baseline time point(5.56±1.22), meaningful time point(5.35±1.26) and validated time point(5.35±1.31) had significant difference, F=3.386,P=0.037. After propensity score matching (PSM) for age and gender, the leukocyte counts at baseline time point had significant between CAS group and healthy controls. Nevertheless, there were no significant differences observed at other time points(Table S1,Figure S2).

## Discussion

In a retrospective cohort of 1141 adults in the Chinese assessed by carotid ultrasound, after CAS first appearance, a retrospective analysis of the previous nearest examination results revealed that CAS patients had higher leukocyte count and LDL-C. While both lab test results provided predictive value, leukocyte count performed significantly better than LDL-C after adjustment for age, gender. Increasingly, both C-IMT and CAP could appear for the first time in newly diagnosed CAS. No transition from C-IMT to CAP was found over a 1.04 years (range:0.58-4.23 years) follow-up observation.

### Prevalence feature of subclinical CAS and the significance of carotid ultrasound detection

CAS was associated with fatty liver^13^ and immune diseases^14^, therefore in this study fatty liver was excluded by liver ultrasound, while immune diseases were excluded by questionnaire. Population aged 50–59 years had the highest percentage change of increased C-IMT and CAP^15^, in our results the CAS group patients mean age after PS matching was 49.6 years, consistent with previous research^16^. C-IMT and CAP do not represent the degree of CAS, even in those with normal C-IMT, mixed or soft plaque was common^17^. While, in our study, not only C-IMT or CAP, but also simultaneous C-IMT and CAP in CAS group were no significant change even compared with the validated follow-up carotid ultrasound. Provided the further evidence that CAS new lesions were found no obvious specificity and no significant changes during subsequent follow-up periods.

### Association between leukocyte counts, LDL-C and CAS

Leukocyte count was important inflammatory marker, while inflammatory participated in both the initiation and perpetuation of atherosclerotic process^18,19^. Moreover, leukocyte counts were significantly associated with HDL-C, LDL-C, TG and TC levels^,20,,21^. Additionally, VLDLR mRNA expressed on peripheral leukocyte independent of the leukocyte subtypes might play an important role in the occurrence of CAS in healthy individuals. And parts of patients characterized by low VLDL-C but high VLDLR mRNA expression in peripheral leukocyte^22^. VLDLR of leukocyte interacts through the fibrin βN-domains(β40-66),and this interaction promotes promoted endothelial permeability and leukocyte transmigration^23^. This pathway is inhibited by the interaction of β15-42 with a putative endothelial receptor^24^. Furthermore, anti-VLDLR monoclonal antibodies (1H10, 1H5) efficiently inhibit leukocyte transmigration induced by fibrin mimetic NDSK-II^25^.

Monocyte count may be a determining factor for CAP but not for C-IMT^26^..But our study showed no significant association between any leukocyte subtypes and the first appearance of CAS. Caucasian individuals under the age of 55, all the arteries deteriorated faster in men than women, and atherosclerotic degeneration began in the abdominal aorta, then the thoracic, coronary, carotid, ascending, and cerebral arteries^27^. Antagonistic interactions between sex and leukocyte count, basophil-to-leukocyte ratio, and platelet counts influenced plaque instability^28^. In our study, after propensity score matching, the clinical characteristics of the CAS group were mainly appearance healthy middle-aged women, with low LDL-C and no significant correlation in leukocyte subtypes. Increasingly, elevated leukocyte count only occurs before the onset of CAS. It had been validated again that the leukocyte count increase could serve as a warning indicator for CAS. The results of this study will promote clinical attention to the risk factors of the above types of populations and strengthen early intervention.

### Strengths and Limitations

To our knowledge, this was the first study to reveal warning indicators for CAS and the onset time after warning. However, our study had certain limitations. First, all participants’ past and family medical histories were self-reported, parts of the histories were validated by the electronic medical records, but we did not have reliable access to all the histories.

Second, the study participants were mostly Han Chinese and above half were women. Third, the study participants were regionally confined. The indicators of systemic inflammation ^29^, monocyte-macrophage^30^, coagulation and fibrinolytic disturbances^31-33^, and endothelial dysfunction^34^were associated with vascular injury. This study was limited by the examination items of the participants, and there were few inflammatory indicators observed during follow-up. All these limitations were unavoidable. In future research, we plan to collaborate with multiple centers to verify the credibility of the conclusion.

## Conclusion

This study results place importance on the timing of CAS early warning and lifestyle interventions. The current guidelines are based solely on management of CAS related to organ damage, nevertheless, lifestyle intervention and medical treatment at this time points were difficult to improve the progression of CAS. Thus, there is a dire need for early screening and management guidelines. Our study pinpointed leukocyte count and LDL-C linked to the onset of CAS in the Chinese physical examination population. Leukocyte count above 5.00*10^9^/L more accurately predict CAS progression than LDL-C above 125.1mg/dl after an average of 1.09 years in middle aged adults. Additionally, leukocyte count no longer increasing after the baseline time point validated again that the leukocyte count increase can serve as a warning indicator better than LDL-C. More importantly, these two indicators are simple to operate, highly credible, making them easily implementable into monitoring indicators. This work and the prospective follow-up studies held the significance first step in early warning and intervention for CAS.

## Data Availability

The ethics panel of Tsinghua Changgung Hospital granted permission for the research (ethics number:24634-6-01). Since the analysis only utilized unidentified data, individual consent was exempted.

## Reference

1. Fuster V, García-Álvarez A, Devesa A, et al. Influence of Subclinical Atherosclerosis Burden and Progression on Mortality. J Am Coll Cardiol. 2024 Oct 8;84(15):1391–1403.

2. Fegers-Wustrow I, Gianos E, Halle M, Yang E. Comparison of American and European Guidelines for Primary Prevention of Cardiovascular Disease: JACC Guideline Comparison. J Am Coll Cardiol. 2022 Apr 5;79(13):1304–1313.

3. Martin SS, Aday AW, Almarzooq ZI, et al; American Heart Association Council on Epidemiology and Prevention Statistics Committee and Stroke Statistics Subcommittee. 2024 Heart Disease and Stroke Statistics: A Report of US and Global Data From the American Heart Association. Circulation. 2024 Feb 20;149(8):e347–e913.

4. Nicolaides AN, Panayiotou AG, Griffin M, Tyllis T, et al. Arterial Ultrasound Testing to Predict Atherosclerotic Cardiovascular Events. J Am Coll Cardiol. 2022 May 24;79(20):1969–1982.

5. Liang LR, Wong ND, Shi P, et al. Cross-sectional and longitudinal association of cigarette smoking with carotid atherosclerosis in Chinese adults. Prev Med. 2009 Aug;49(1):62–7.

6. Inaba Y, Chen JA, Bergmann SR. Carotid plaque, compared with carotid intima-media thickness, more accurately predicts coronary artery disease events: a meta-analysis. Atherosclerosis 220(1):128–133.

7. Martin SS, Aday AW, Almarzooq ZI, et al. American Heart Association Council on Epidemiology and Prevention Statistics Committee and Stroke Statistics Subcommittee. 2024 Heart Disease and Stroke Statistics: A Report of US and Global Data From the American Heart Association. Circulation. 2024 Feb 20;149(8):e347–e913.

8. Shai I, Spence JD, Schwarzfuchs D,et al; DIRECT Group. Dietary intervention to reverse carotid atherosclerosis. Circulation. 2010 Mar 16;121(10):1200–8.

9. Crouse JR 3rd, Raichlen JS, Riley WA, et al; METEOR Study Group. Effect of rosuvastatin on progression of carotid intima-media thickness in low-risk individuals with subclinical atherosclerosis: the METEOR Trial. JAMA. 2007 Mar 28;297(12):1344–53.

10. Yuan S, Li L, Pu T, et al. The relationship between NLR, LDL-C/HDL-C, NHR and coronary artery disease. PLoS One. 2024 Jul 10;19(7):e0290805.

11. Nicolaides AN, Panayiotou AG, Griffin M, et al. Arterial Ultrasound Testing to Predict Atherosclerotic Cardiovascular Events. J Am Coll Cardiol. 2022 May 24;79(20):1969–1982.

12. Kabłak-Ziembicka A, Przewłocki T. Clinical Significance of Carotid Intima-Media Complex and Carotid Plaque Assessment by Ultrasound for the Prediction of Adverse Cardiovascular Events in Primary and Secondary Care Patients. J Clin Med. 2021 Oct 9;10(20):4628.

13. Tang ASP, Chan KE, Quek J, et al. Non-alcoholic fatty liver disease increases risk of carotid atherosclerosis and ischemic stroke: An updated meta-analysis with 135,602 individuals. Clin Mol Hepatol. 2022 Jul;28(3):483–496.

14. Rodriguez T, Lehker A, Mikhailidis DP, et al. Carotid Artery Pathology in Inflammatory Diseases. Am J Med Sci. 2022 Mar;363(3):209–217.

15. Song P, Fang Z, Wang H, et al. Global and regional prevalence, burden, and risk factors for carotid atherosclerosis: a systematic review, meta-analysis, and modelling study. Lancet Glob Health. 2020 May;8(5):e721–e729.

16. Sebastian SA, Co EL, Tidd-Johnson A, et al. Usefulness of Carotid Ultrasound Screening in Primary Cardiovascular Prevention: A Systematic Review. Curr Probl Cardiol. 2024 Jan;49(1 Pt C):102147.

17. Boulos NM, Gardin JM, Malik S, et al. Carotid Plaque Characterization, Stenosis, and Intima-Media Thickness According to Age and Gender in a Large Registry Cohort. Am J Cardiol. 2016 Apr 1;117(7):1185–91.

18. Liao M, Liu L, Bai L, et al. Correlation between novel inflammatory markers and carotid atherosclerosis: A retrospective case-control study. PLoS One. 2024 May 29;19(5):e0303869.

19. Huang ZS, Jeng JS, Wang CH, et al. Correlations between peripheral differential leukocyte counts and carotid atherosclerosis in non-smokers. Atherosclerosis. 2001 Oct;158(2):431–6.

20. Liu Y, Kong X, Wang W, et al. Association of peripheral differential leukocyte counts with dyslipidemia risk in Chinese patients with hypertension: insight from the China Stroke Primary Prevention Trial. J Lipid Res. 2017 Jan;58(1):256–266.

21. Hu W, Zhang P, Su Q, et al. Peripheral leukocyte counts vary with lipid levels, age and sex in subjects from the healthy population. Atherosclerosis. 2020 Sep;308:15–21.

22. Zhao F, Qi Y, Liu J, et al. Low Very low-Density Lipoprotein Cholesterol but High Very low-Density Lipoprotein Receptor mRNA Expression in Peripheral White Blood Cells: An Atherogenic Phenotype for Atherosclerosis in a Community-Based Population. EBioMedicine. 2017 Nov;25:136–142.

23. Yakovlev S, Medved L. Dual functions of the fibrin βN-domains in the VLDL receptor-dependent pathway of transendothelial migration of leukocytes. Thromb Res. 2022 Jun;214:1–7.

24. Yakovlev S, Strickland DK, Medved L. Current View on the Molecular Mechanisms Underlying Fibrin(ogen)-Dependent Inflammation. Thromb Haemost. 2022 Nov;122(11):1858–1868.

25. Yakovlev S, Belkin AM, Chen L, et al. Anti-VLDL receptor monoclonal antibodies inhibit fibrin-VLDL receptor interaction and reduce fibrin-dependent leukocyte transmigration. Thromb Haemost. 2016 Nov 30;116(6):1122–1130.

26. Johnsen SH, Fosse E, Joakimsen O, et al. Monocyte count is a predictor of novel plaque formation: a 7-year follow-up study of 2610 persons without carotid plaque at baseline the Tromsø Study. Stroke. 2005 Apr;36(4):715–9.

27. Jakab AE, Bukva M, Maróti Z, et al. The ASAP study: association of atherosclerosis with pathobiology in a caucasian cohort-a study of 3400 autopsy reports. Sci Rep. 2024 Oct 24;14(1):25179.

28. Gasbarrino K, Zheng H, Daskalopoulou SS. Circulating Sex-Specific Markers of Plaque Instability in Women and Men With Severe Carotid Atherosclerosis. Stroke. 2024 Feb;55(2):269–277.

29. Song JE, Hwang JI, Ko HJ, et al. Exploring the Correlation between Systemic Inflammatory Markers and Carotid Atherosclerosis Indices in Middle-Aged Adults: A Cross-Sectional Study. J Cardiovasc Dev Dis. 2024 Feb 21;11(3):73.

30. Winkels H, Ehinger E, Vassallo M, et al. Atlas of the Immune Cell Repertoire in Mouse Atherosclerosis Defined by Single-Cell RNA-Sequencing and Mass Cytometry. Circ Res. 2018 Jun 8;122(12):1675–1688.

31. Gravholt CH, Mortensen KH, Andersen NH, et al. Coagulation and fibrinolytic disturbances are related to carotid intima thickness and arterial blood pressure in Turner syndrome. Clin Endocrinol (Oxf). 2012 May;76(5):649–56.

32. Shi P, Zheng W, Zhou J, Han N, et al. Effects of MaiLiuPian on carotid thrombosis in rats and acute pulmonary embolism in mice and its antithrombotic mechanism. J Food Biochem. 2022 Jul;46(7):e14143.

33. Wang X, Li S, Liu C, et al. High expression of PLA2G2A in fibroblasts plays a crucial role in the early progression of carotid atherosclerosis. J Transl Med. 2024 Oct 24;22(1):967.

34. Hsu PL, Chen JS, Wang CY, et al. Shear-Induced CCN1 Promotes Atheroprone Endothelial Phenotypes and Atherosclerosis. Circulation. 2019 Jun 18;139(25):2877–2891.

